# Cohort profile: A community-based prospective cohort study of Alzheimer’s disease and related dementias in the Democratic Republic of Congo

**DOI:** 10.1101/2025.09.23.25336499

**Authors:** Jean Ikanga, Caterina Obenauf, Saranya Sundaram Patel, Megan Schwinne, Guy Gikelekele, Emmanuel Epenge, Japhet Magolu Potshi, Topina Tomadia, Julie Kazadi, Immaculee Kavugho, Giovanni Luamba Santolini, Francine Manyonga Sabowa, Brenda Matungu Musewu, Elipsis Mawisa, Joseph Phuati Tsangu, Francis Muena Kondo Beya, Samuel Mampunza, Lelo Mananga, Justin Bukabau, Charlotte E. Teunissen, Thomas Karikari, Alden L. Gross, Julio C. Rojas, Alvaro Alonso

## Abstract

**INTRODUCTION:** The *Étude du Vieillissement Cognitif et de Démence en République Démocratique du Congo* (Study of Cognitive Aging and Dementia in the Democratic Republic of Congo, EVCD-RDC) was launched in 2024 to characterize cultural, biological, and environmental risk factors of cognitive aging and dementia in Kinshasa.

**METHODS:** The study is enrolling 800 adults aged ≥65 years across four diagnostic groups (cognitively unimpaired, mild cognitive impairment, Alzheimer’s disease [AD], and other dementias). Recruitment occurs through community and clinical settings. Assessments include neuropsychological testing, neurological examination, social determinants, biospecimen collection, and air pollution exposure, with follow-up every two years.

**RESULTS:** To date, 506 participants have been enrolled (mean age: 73.4 years; 59.3% women). Dementia cases had greater prevalence of hypertension and other comorbidities, more neuropsychiatric symptoms, higher rates of smoking and alcohol use, lower body weight, reduced educational attainment and resilience, and increased exposure to air pollution, war, traumatic experiences, and poverty. The internal consistency of cognitive measures ranged from 0.44 to 0.91.

**DISCUSSION:** EVCD-RDC establishes the first large-scale dementia cohort in French-speaking Sub-Saharan Africa, providing critical infrastructure and insights into risk factors for Alzheimer’s disease and related dementias.

## 1. INTRODUCTION

Cognitive aging and dementia are emerging global public health concerns due to aging populations worldwide. Recent estimates indicate that more than 50 million people are affected by dementia globally; this figure is projected to nearly triple by 2050, disproportionately impacting low- and middle-income countries mainly due to population growth and aging.^1^ The increasing burden of dementia and cognitive aging issues not only overwhelms healthcare systems and social services, but it also places a financial and emotional burden on individuals, families, and communities, with global costs beyond $1 trillion annually.^2^

As the aging population increases globally, the rising incidence of dementia necessitates a thorough investigation of its evolving epidemiology and treatment approaches. Longitudinal cohort studies have proven particularly valuable for this purpose, as they allow for monitoring of the prevalence, incidence, and progression of cognitive impairment over time.^3,4^ Such studies also enable the identification of risk and protective factors while capturing inter-individual variability of cognitive aging.^3,4^ Moreover, cohort studies are essential for characterizing cognitive decline across populations and cultures.^4^ As dementia risk and presentation may vary by cultural, genetic, and socioeconomic contexts, it is critical to examine cognitive aging across a variety of global settings.

In high-income countries, well-established programs such as the Framingham Heart Study in the United States^5^ and the Whitehall II Study in the United Kingdom^6^ have provided critical insights into the risk factors and progression of cognitive aging. Cohort studies in middle- and low-income countries, including the 10/66 Dementia Research Group studies in Asia and Latin America^7^ and the Brazilian Longitudinal Study of Aging (ELSI-Brazil)^8^ are beginning to reveal unique epidemiologic patterns driven by differing socioeconomic and lifestyle factors. In sub-Saharan Africa, English-speaking countries such as Nigeria and South Africa have initiated cohort studies to explore dementia and aging.^9,10^ However, research in French-speaking African countries remains limited, with few prospective cohort studies conducted in these contexts.^11^ Prevalence and risk factors of dementia have been examined in the French-speaking Republic of Congo,^12^ Central African Republic,^13^ and Benin.^14^ In the Democratic Republic of the Congo (DRC) specifically, there have also been cross-sectional studies,^15^ but no longitudinal cohort studies, reflecting complex challenges such as linguistic and cultural barriers to research.^16^

To aid in addressing this gap, we have established the *Étude du Vieillissement Cognitif et de Démence en République Démocratique du Congo* (EVCD-RDC; Study of Cognitive Aging and Dementia in the Democratic Republic of the Congo), which is recruiting and prospectively following a representative sample of older adults in the French-speaking DRC to investigate the clinical course, biomarkers, and neuropsychiatric symptoms of cognitive aging and dementia within this understudied population. In this cohort profile paper, we describe the origins, study design, and key enrollment characteristics of this unique cohort and present a preliminary overview of our findings. By doing so, we aim to inform prevention and treatment programs not only in the DRC but also in similar contexts across French-speaking Africa.

## 2. SUBJECTS/MATERIALS AND METHODS

### 2.1 Cohort description

#### 2.1.1 Cohort setting, objectives, and design

The EVCD-RDC is a prospective, longitudinal cohort study based in Kinshasa, the capital of the Democratic Republic of Congo (the second-largest country in Africa). The study will soon expand to other major cities, including Lubumbashi, Mbuji-Mayi, Kisangani, and Mbandaka.

The key objectives of this cohort study are to: (1) estimate prevalence, incidence, and risk factors for cognitive aging and dementia in this population; (2) track the progression of cognitive aging and dementia by monitoring functional, behavioral, and cognitive symptoms, as well as neurodegenerative biomarkers over time; (3) validate neurodegenerative biomarkers and reference cutoffs for early dementia diagnosis, disease monitoring, and treatment response; (4) develop and refine diagnostic criteria, predictive models, and prognostic tools in this new population; and (6) assess the needs of individuals with dementia and the burden on caregivers and family members to inform strategies for caregiving and provide evidence-based recommendations for healthcare policy, practice, and guidelines.

### 2.2 Overall study design

The study design consists of the following steps: (1) Recruitment & Enrollment: Participants are recruited from churches, senior associations, military camps, hospitals, and private clinics. In collaboration with community leaders, the PI shared and discussed a written announcement with the community. The announcement outlined the inclusion and exclusion criteria, emphasized the importance of the research for the country and medical community, and highlighted its relevance to the older adult population. Volunteers were then enrolled to be screened by the research team. (2) Screening & Evaluation: Screening tests, questionnaires, and neurocognitive assessments are conducted at the recruitment sites. (3) Sample Collection & Neurological Evaluation: Biological samples (cerebrospinal fluid, blood, urine, saliva, and feces) and neurological assessments are conducted at Centre Medical de Kinshasa (CMK) to minimize misconceptions about witchcraft when specimen collection occurs outside clinical settings. (4) Cohort Follow-Up: Follow-ups—including neurocognitive tests, neurological evaluations, and specimen collection—will take place every two years. The initial recruitment, enrollment, screening, evaluation, and sample collection phase took place over 13 months (from August 2024 to September 2025). The second re-evaluation will begin in August 2026, contingent on funding availability. Figure 1 illustrates the recruitment, screening, and sample collection processes for the initial evaluation.

**Figure 1.**
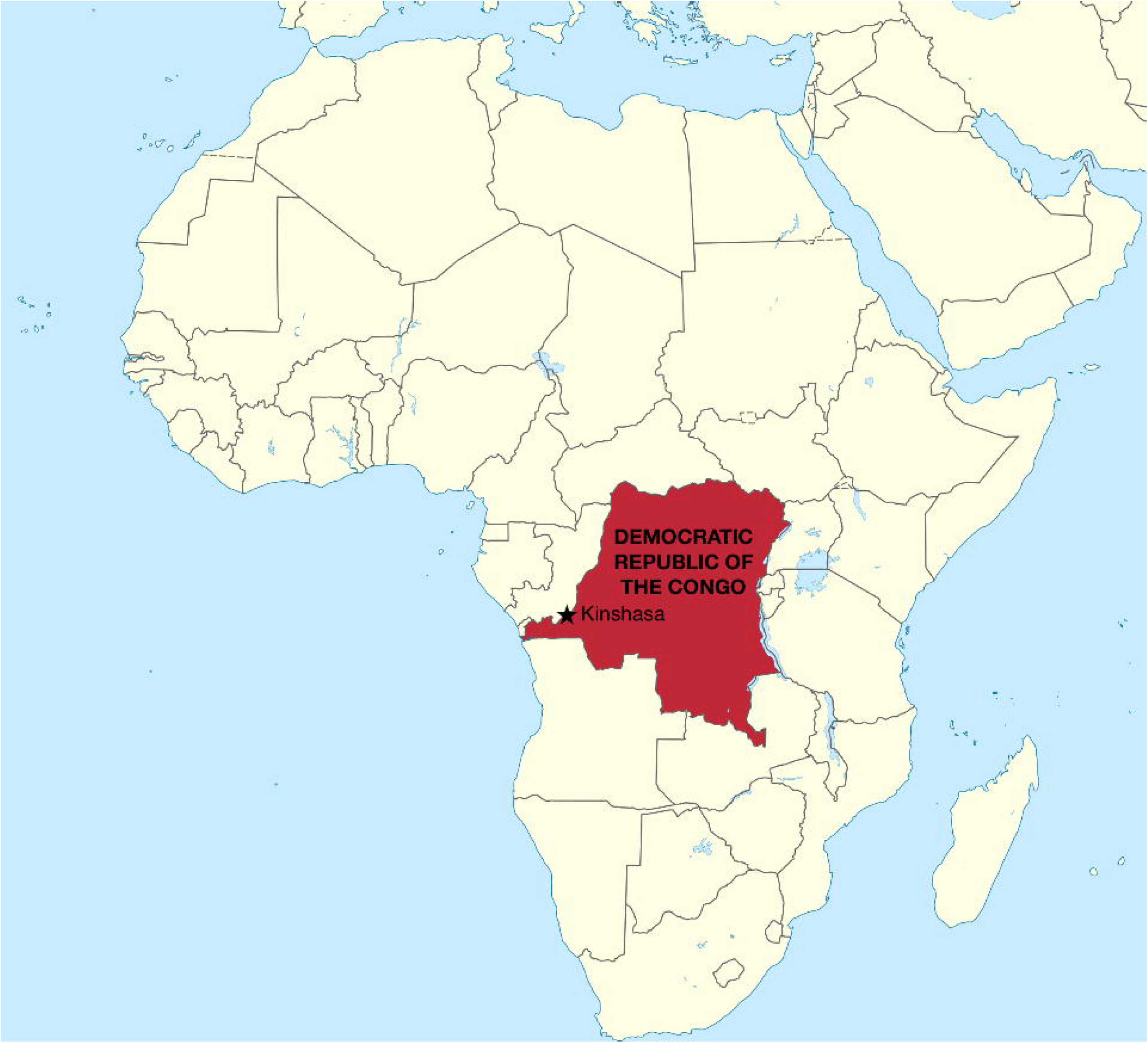
Map of the Democratic Republic of the Congo (DRC) highlighting Kinshasa, the capital and primary study site for the Étude du Vieillissement Cognitif et de Démence en République Démocratique du Congo (EVCD-RDC).

#### 2.2.1 Cohort Eligibility

Eligibility was determined during recruitment and screening visits (see Figure 1). To be eligible for inclusion in this open cohort, individuals must: (1) be Congolese citizens residing in the Democratic Republic of Congo; (2) be 65 years or older; (3) provide a valid home address, phone number, and the contact information of a caregiver or informant; (4) demonstrate willingness to participate in longitudinal study activities, including initial enrollment, follow-up visits, and baseline specimen collection; (5) have the ability to provide informed consent, either independently or via a caregiver; (6) be fluent in Lingala or French; and (7) have no prior neurodevelopmental, psychiatric, or neurological disorders affecting the central nervous system. Individuals with significant vision or auditory impairments were excluded.

### 2.3 Study measurements

The measures included the Uniform Data Set - Version 3 (UDS-3), screening procedures, neurocognitive tests—such as the African Neuropsychology Battery - Short Version (ANB-SV)—intelligence tests, and other questionnaires (see Table 1). These tests were derived from previous dementia cohort studies^17,18^ to harmonize results across cohorts and align with international studies. Additional measures, including cognitive tests and questionnaires, were selected to address specific issues related to aging in Sub-Saharan Africa and Congo, such as resilience, poverty, illiteracy, air pollution, and cardiovascular health. We calculated the total scores from the resilience and poverty questionnaires to derive the respective resilience and poverty indices. These instruments were adapted to the Sub-Saharan African context (see Table 1), translated into French and Lingala (a Congolese language), and pre-piloted before being implemented in the study. Participants underwent a comprehensive neurological evaluation, which included an assessment of cranial nerves, gait and balance, motor coordination, archaic reflexes, overall muscle strength, segmental muscle strength, muscle tone, osteotendinous reflexes, and superficial and deep sensitivity. The follow-up assessments are outlined in Table 1.

### 2.4 Clinical adjudication process

Participants were classified as being cognitively unimpaired (CU), or having mild cognitive impairment (MCI), suspected Alzheimer’s disease (susp AD), or another form of dementia (OD). This classification was conducted by a team consisting of a neuropsychologist (JI), neurologist (EE or PJ), psychiatrist (GG), and internal medicine physician (TT). The clinical adjudication process involved two stages: (1) determining whether participants had cognitive deficits (CU or not CU), and (2) identifying the possible etiology of the cognitive deficits based on the number and severity of impairments, medical history, neurological evaluation, and lifestyle factors. Following the clinical adjudication algorithm outlined by Manly and colleagues,^19^ participants who exhibited no cognitive impairment in any domain and demonstrated normal functioning—according to IDEA—were classified as CU. Those with deficits in one cognitive domain but preserved functional ability, as defined by IDEA, were classified as MCI. Participants who showed impairment in at least two cognitive domains (scores >1.5 standard deviations below the mean) and functional impairment per IDEA criteria were categorized as having dementia. Participants who reported a progressive decline and exhibited severe, widespread deficits—primarily in learning and memory—were classified as suspected Alzheimer’s disease (AD). In contrast, individuals with a stepwise decline, cognitive impairments linked to other medical conditions, fluctuating cognitive performance, or marked personality changes were categorized as having other forms of dementia (see Figure 2).

**Figure 2.**
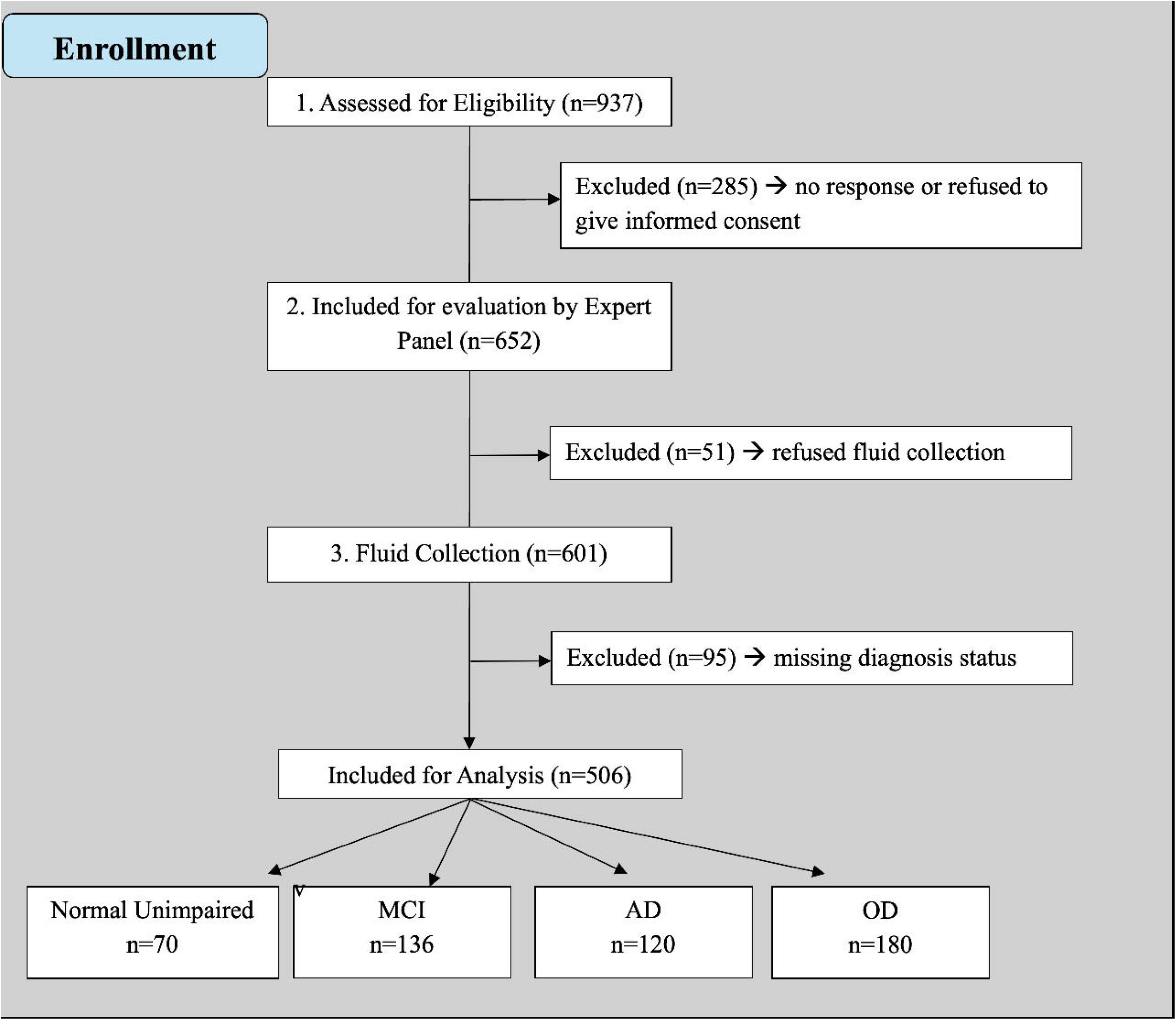
Clinical adjudication process for classifying participants as cognitively unimpaired (CU), mild cognitive impairment (MCI), suspected Alzheimer’s disease (AD), or other dementia (OD). The process involved two steps: (1) determining presence of cognitive deficits, and (2) evaluating possible etiology based on neuropsychological performance, functional status (IDEA), medical history, and neurological/psychiatric evaluations. Classifications were adjudicated by a multidisciplinary team (neuropsychology, neurology, psychiatry, and internal medicine).

**Figure 3.**
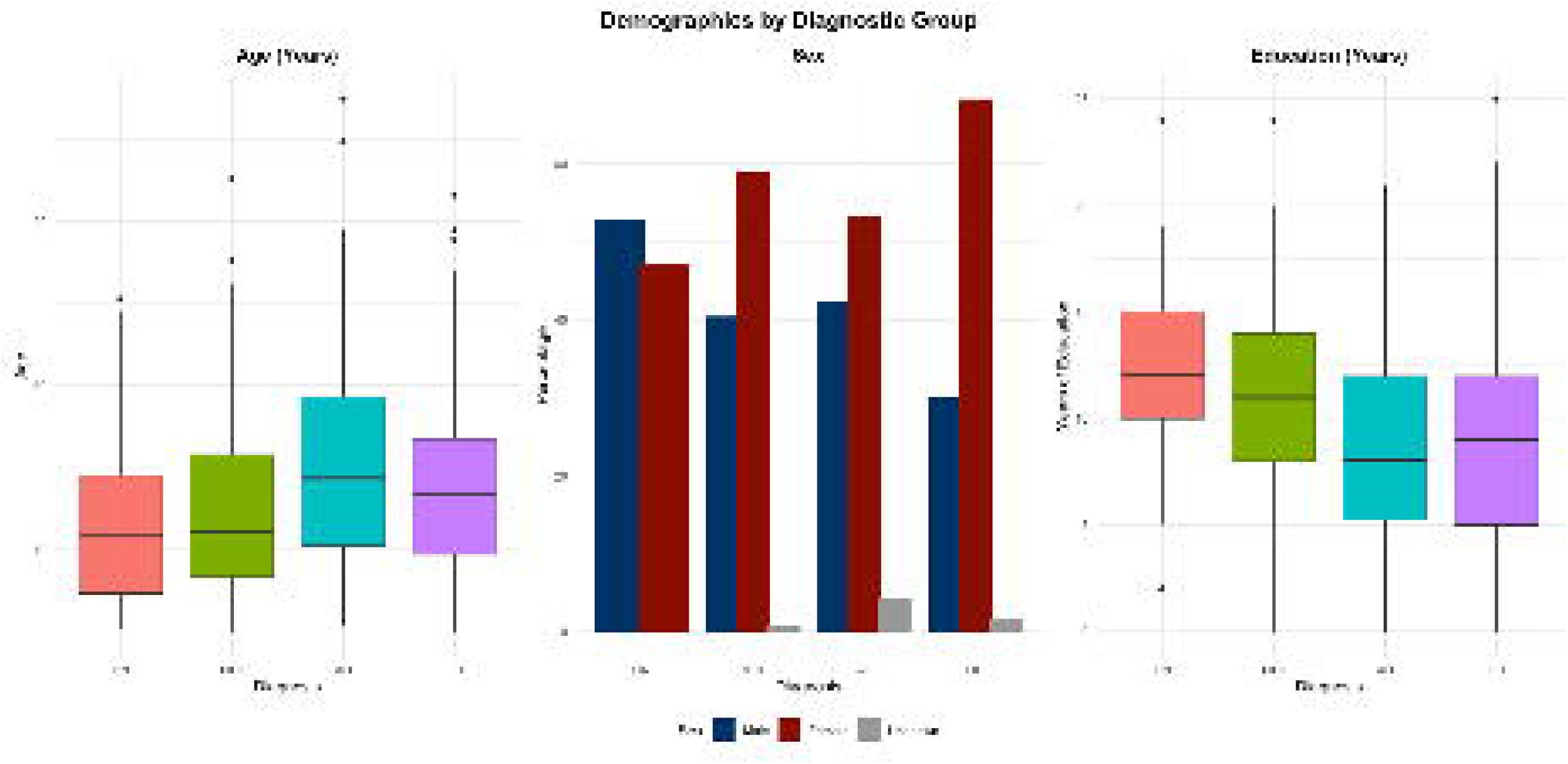
Distribution of mild cognitive impairment (MCI), Alzheimer’s disease (AD), and other dementias (OD) by gender, age, and years of education among enrolled EVCD-RDC participants.

To determine the predicted score of the participant, we applied a linear regression model using scores from screening, functional, cognitive, and intelligence tests. This model examined the relationship between predictors and cognitive outcomes. In specifying the model, outcome variables included the screening tests, cognitive tests, intelligence assessments, and functional tests, while predictor variables consisted of demographic factors (age, gender, and years of education). Our regression equation was as follows:

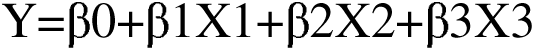

where:

- Y represents screening test scores, cognitive test scores, functional test scores, or intelligence test scores,
- X1 = education,
- X2 = age,
- X3 = gender.

The linear regression equation was Y=β0+β1(education)+β2(age)+β3(gender). The coefficients β0, β1, β2, and β3 were obtained based on our previous study. ^15^Our model was evaluated using the R-squared value, and our clinical interpretations were guided by the context of clinical practice in Congo. We considered the strength and significance of associations between predictors and cognitive outcomes, as well as the model’s ability to predict cognitive decline or dementia.

To assess participant impairment in the test and determine how many standard deviations a participant’s predicted score deviates from the population mean, we calculate the z-score using the test scores from our CU in a previous study^15^:

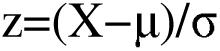

Where:

- X was the participant’s predicted score based on the linear regression equation.
- μ represented the population mean.
- σ denoted the population standard deviation.

#### 2.4.1 Collection of biological fluid

Various biological fluids were collected from participants following international standards at the Centre Médical de Kinshasa (CMK) by trained laboratory technicians. Cerebrospinal fluid (CSF) was collected from 200 participants (50 in each category), while blood (for plasma, serum, PAXgene RNA, and DNA), urine, saliva, and feces were collected from all participants. CSF was obtained using a sterile, single-use polypropylene tube. Non-hemolyzed blood was drawn via venipuncture. The blood collection process included:

- A 10cc dry tube without anticoagulant for serum.
- Three EDTA tubes with anticoagulant: one 10cc tube for plasma and two 4cc tubes for DNA.
- An additional 2.5ml of blood was drawn into a PAXgene Blood RNA tube containing an intracellular RNA stabilizer.

Saliva was collected in a Sarstedt Salivette tube by placing a cotton roll in the participant’s mouth and instructing them to saturate it with saliva. After two minutes, the saliva-soaked cotton roll was removed and placed into a collection tube. Urine was collected in a Sarstedt cat bottle, and participants were advised not to drink water or other liquids prior to collection. Fecal samples were brought from participants’ homes.

##### Centrifugation and aliquoting

Blood (for serum and plasma) and CSF in the same tubes of collection and urine in conic sterile tube underwent centrifugation and aliquoting. Blood collected without anticoagulants was left at room temperature for at least 30 minutes before processing.

The centrifugation steps were as follows:

- Blood, CSF, and urine were centrifuged at 4000 rpm for 10 minutes.
- Processed specimens were placed on a rack and pipetted into 0.5 ml (500 µL) aliquots to obtain serum and plasma.
- Urine was transferred into sterile 6 ml conical tubes before centrifugation.
- PAXgene Blood RNA, saliva, and feces were not centrifuged.

#### 2.4.2 Sample storage

CSF, plasma, serum, and urine aliquots were stored directly in cryotubes at −20°C, then transferred to −80°C to ensure biomarker precision and reliability. Saliva was stored immediately after collection in its original tube. Feces and PAXgene Blood RNA samples were first stored at −20°C for 24 hours, then transferred to −80°C for long-term preservation.

#### 2.4.3 Pre-analytic and biobanking

These phases were critical in our cohort study to ensure: (1) sample quality—proper collection, handling, and storage of biological specimens; (2) reduction of errors and variability in sample collection, ensuring accurate and reliable biomarker measurements; (3) minimization of false-positive and false-negative results; (4) laboratory efficiency, optimizing workflow and specimen processing; and (5) regulatory compliance, adhering to strict protocols for sample collection, handling, and storage. To maintain high standards, our laboratory technicians identified and labeled each sample based on the participant’s ID, type of fluid, classification from clinical adjudication, and date of collection. To ensure proper sample collection, handling, and storage, we meticulously documented the type of biological fluid, the tube used for collection, centrifugation conditions, and storage protocols. Additionally, we recorded the duration between collection and centrifugation, as well as the time from centrifugation to storage at −20°C and −80°C. To enhance our data management, sample integrity, medical research, and personalized medicine in Congo, we acquired a −80°C freezer to immediately store collected samples and preserve their quality.

#### 2.4.4 Estimating kidney function

The estimated Glomerular Filtration Rate (eGFR) is commonly used as a measure of kidney function. Previous studies on neurodegenerative plasma biomarkers have highlighted the critical role of eGFR, as kidney function can influence the concentration of certain biomarkers (e.g., amyloid-beta and tau) in the bloodstream, as well as their clearance. Therefore, eGFR is considered a potential confounding factor in biomarker studies.^20^ To assess kidney function in our sample, we calculated eGFR using serum creatinine levels. Bukabau and colleagues^21,22^ found that commonly used eGFR equations [Chronic Kidney Disease Epidemiology Collaboration (CKD-EPI), the European Kidney Function Consortium (EKFC), and the revised Lund-Malmo Revised (r-LMR)], even those adjusted for race, were not applicable to SSA populations. Based on their previous findings, we calculated the estimated glomerular filtration rate (eGFR) using the Modification of Diet in Renal Disease (MDRD) formula, as our serum creatinine measurements were not calibrated. After adjusting for demographic factors such as age and gender, the eGFR was calculated using the following formula:

eGFR (MDRD) = 175 × (Serum Creatinine) ^–1.154 × (Age)^–0.203 × (0.742 if female). This formula is widely used to monitor kidney function over time and to help diagnose chronic kidney disease (CKD).^21,22^

#### 2.4.5 Air pollution

Research has shown that air pollution can trigger neuroinflammation, oxidative stress, endothelial dysfunction, and epigenetic changes.^23–25^ To assess local air quality, we are using an ATMO Tube PRO device to collect data on the following pollutants and environmental parameters: PM1, PM2.5, PM10, barometric pressure, volatile organic compounds (VOCs), temperature, humidity, and altitude. Data collectors were trained by a team expert on how to use the ATmoTube PRO device. Each collector wore the device for one hour inside the participant’s residence, measuring air pollutants every 15 minutes. A total of four readings were recorded. These readings were summed and averaged to estimate the air pollution level for each participant.

### 2.5 Data management

Study data were collected and managed using REDCap (Research Electronic Data Capture), a secure, web-based application designed to support research data collection.^26,27^ All study variables, including sociodemographic characteristics, clinical indicators, validated psychological and cognitive measures, and biological samples, were entered directly into REDCap by trained research personnel. Data were monitored regularly for completeness, consistency, and quality. Built-in REDCap data quality rules were used to flag potential data entry errors, which were reviewed and resolved by the data management team. Duplicate entries were checked using participant IDs. After data collection was complete, a final review of data quality was conducted prior to statistical analyses.

The de-identified dataset was exported from REDCap into IBM SPSS Statistics version 29.0 for analysis. Prior to statistical analyses, variables were recoded as needed (e.g., reverse scoring), and total scores were calculated for relevant scales according to established scoring guidelines. Missing data were evaluated descriptively to assess patterns and the extent of missingness across variables. Most missing data resulted from participants not being administered all measures due to time constraints or technical difficulties during data collection. For variables with minimal missingness (<5%), pairwise deletion was used.

### 2.6 Cohort profile analysis

Descriptive analyses, stratified by neurological status, were conducted using means and standard deviations for continuous variables, and frequencies with column percentages for categorical variables. When examining cohort characteristics with neurological status as the outcome, multinomial logistic regression models were fitted, adjusting for age, sex, and education. Likelihood ratio tests (LRT) were performed by comparing the full model to a reduced model with the variable of interest removed to assess its association with neurological status. When evaluating cognitive profiles, cumulative disease assessments, and laboratory metrics with neurological status as the predictor, analysis of variance (ANOVA) models were applied, adjusting for age, sex, and education. For dichotomous outcomes, logistic regression models were used, also adjusting for age, sex, and education, with p-values obtained via LRT. Tukey’s post-hoc tests were conducted for all pairwise comparisons, with the neurological status of *cognitively unimpaired* as the reference group. Statistical significance was defined as *p* < 0.05. All analyses were performed using R (version 5).

## 3. RESULTS

### 3.1 Characteristics of the cohorts

Table 2 presents the sociodemographic characteristics of the cohort, categorized by neurological status. This analysis includes 506 participants with a mean age of 73.4 years (SD = 5.9), of whom 59.3% are women. The average educational attainment is 9.7 years (SD = 4.8), with participants in the cognitively unimpaired (CU) and mild cognitive impairment (MCI) groups having significantly more years of education than those with suspected dementia (Alzheimer’s disease [AD] or other dementias [OD]). The cohort is predominantly Bantu (98%), with a small representation of Nilotic and Sudanic individuals, and no pygmy participants. Regarding marital status, individuals with suspected dementia were more likely to be divorced, whereas those in the CU and MCI groups were more likely to be married. All participants lived with either a spouse or a relative. While most participants lived independently, those with suspected dementia required more assistance with complex daily activities compared to CU and MCI groups. Nearly all participants resided in non-assisted living homes (see Table 2).

### 3.2 Medical history of the cohort

Table 3 summarizes the medical history of the cohort. While the average weight of CU was 78.3 kg, participants with suspected dementia were significantly underweight (68.6 kg for AD and 64.0 kg for OD). The mean BMI was 25.9 kg/m^2^, with lower values observed in the suspected dementia groups. Most participants were hypertensive; 64.9% were aware of their condition, 30.8% were taking antihypertensive medications, and 31.7% had controlled hypertension. Neurological evaluations revealed abnormalities in 21% of the cohort, with the OD group showing the highest rate (31%). Based on cumulative disease assessments, the most reported conditions were eye, ear, nose, and throat disorders (38%); musculoskeletal issues (21%), vascular diseases (17%), and upper gastrointestinal disorders (12%). Creatinine levels were consistent across all four groups, with a mean of 0.90 mg/dL and a median of 0.85 mg/dL. Estimated glomerular filtration rate (eGFR), calculated using the MDRD equation, showed no significant differences between neurological status groups. The mean eGFR ranged from 69.7 to 74.4 mL/min/1.73m², which falls below the normal range (>90 mL/min/1.73m²).

### 3.3 Neuropsychiatric profile of the cohort

Table 4 presents neuropsychiatric symptoms and questionnaire results. Participants with suspected dementia exhibited a higher frequency of neuropsychiatric symptoms (NPS) compared to those in the CU and MCI groups. Nearly half of the participants (44%) had experienced war, and 59 % reported traumatic events within the past 10 years. Only 12% were tobacco smokers, and 21% consumed alcohol about once a month. Sleep-related issues were reported by 31% of the sample, including sleep apnea, sleep disorders, or insomnia (see Table 4). The most common NPS were depression (26%), loss of appetite (21%), irritability (19%) and aggressiveness (19%), with higher prevalence in AD that other groups (see Figure 4). Participants with suspected dementia also had a more adverse profile based on social determinants of health (SDOH) compared to CU and MCI groups. Although the overall cohort had a high resiliency score (mean = 19.9), the suspected dementia group scored lower (18.0) than CU (21.3). Additionally, the suspected dementia group had higher poverty scores (6.8) compared to CU (4.8) and MCI (5.8).

**Figure 4.**
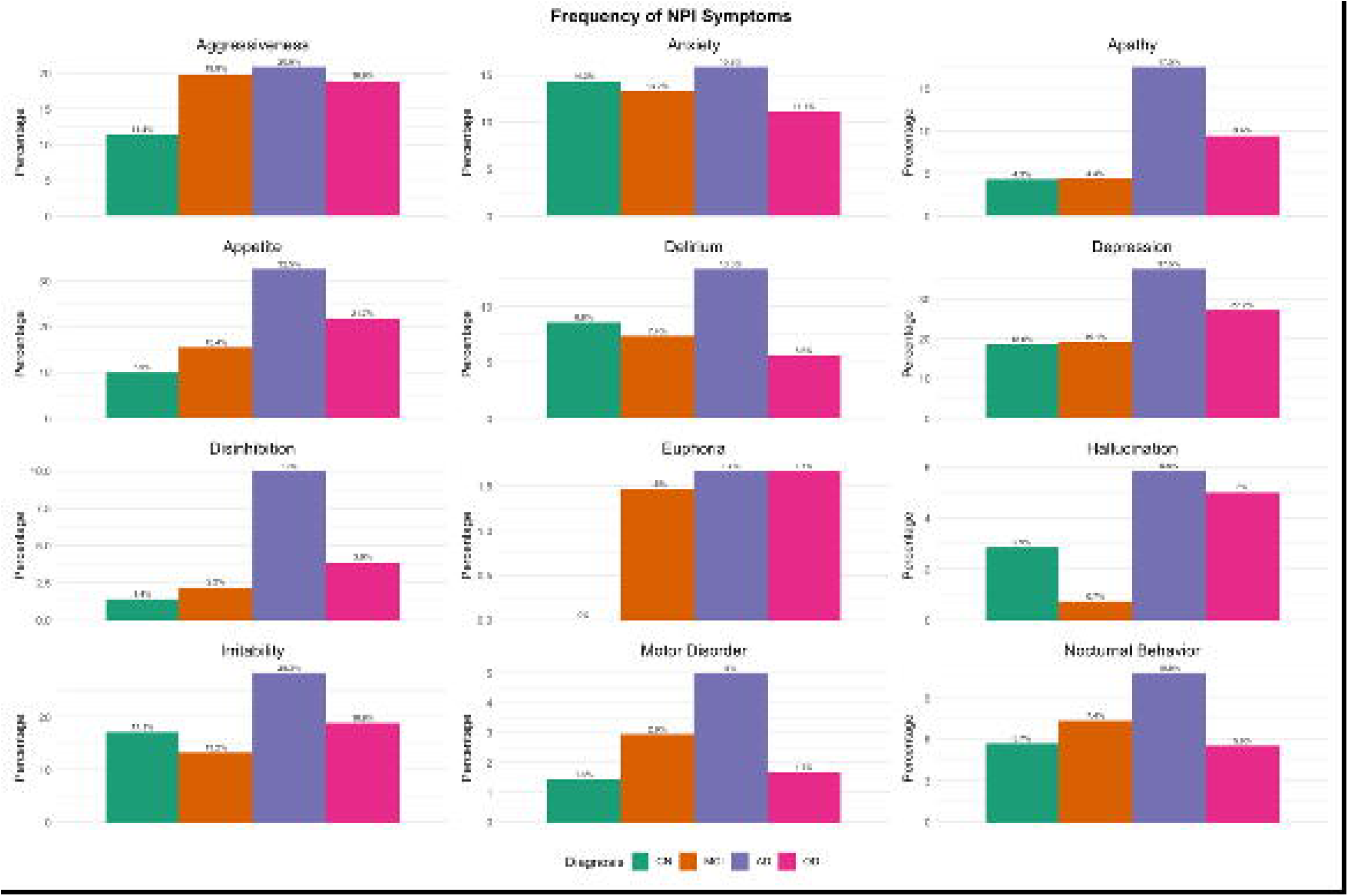
Frequency of neuropsychiatric symptoms as measured by the Neuropsychiatric Inventory (NPI) among EVCD-RDC participants, stratified by diagnostic group.

### 3.4 Cognitive profile of the cohort

Table 5 summarizes the psychometric properties (internal consistency) of screening and cognitive tests. The internal consistency of almost all the screening and cognitive tests, as measured by Cronbach’s alpha, was between good to excellent reliability. Only two tests, the African Visuospatial Memory Test and the African Proverb Test, had acceptable reliability. The internal consistency of the African Emotion Test was questionable (.441).

Table 6 presents the screening and neuropsychological test results. Significant differences were found in mean scores across all screening and cognitive assessments (p = 0.001), with the CU group consistently outperforming the MCI, AD, and OD groups.

### 3.5 Air pollution

Table 7 and Figure 5 depict air pollution levels in Kinshasa. Concentrations of PM1, PM2.5, and PM10 range from 2.8 to 670 µg/m³ (Median = 70 µg/m³), 2.5 to 691.3 µg/m³ (Median = 75 µg/m³), and 5.6 to 692.3 µg/m³ (Median = 76 µg/m³), respectively. No statistically significant differences were observed among the groups for any of the air pollutants.

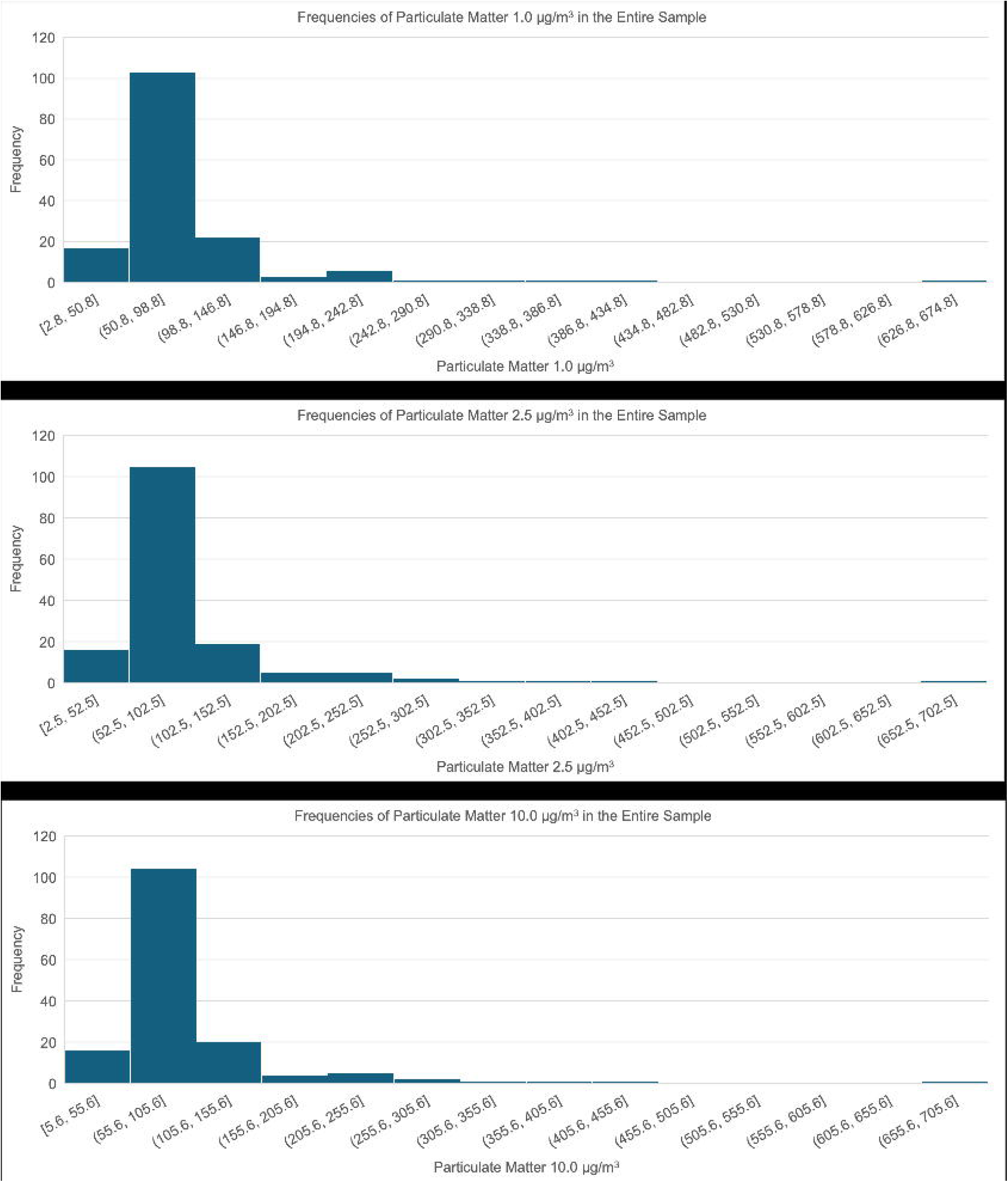

## DISCUSSION

This longitudinal, community-based cohort study has enrolled, as of the date of analysis, 506 individuals from 21 of the 24 quarters of Kinshasa, the capital of the Democratic Republic of Congo (DRC). The cohort is geographically, socio-demographically, and linguistically diverse, and includes participants across the main neurological statuses: Cognitively unimpaired (CU), Mild Cognitive Impairment (MCI), and dementia—either Alzheimer’s Disease (AD) or Other Dementia (OD). In terms of ethnicity, the cohort reflects the demographic composition of Kinshasa, which is approximately 80% Bantu, with smaller representations of Sudanic, Nilotic, and Pygmy groups.^28^

Demographically, this cohort is older (mean age: 73.4 years) than some previous studies, such as the Health and Aging in Africa: Longitudinal Study of an INDEPTH Community (HAALSI), where the average age was around 61 years, with a focus on individuals aged 40 and older.^29^ As seen in many aging and dementia cohorts worldwide (e.g., HAALSI, where 53.6% of 5,059 participants were women), women are more represented, likely due to their longer life expectancy, greater willingness to participate in research, and higher healthcare-seeking behavior. ^29^ Additionally, recruitment through churches may have contributed to this gender imbalance, as women tend to attend church more frequently than men.

As emphasized by the 2024 Lancet Commission, low education is a known risk factor for dementia.^3^ In this cohort, individuals with suspected dementia had lower educational attainment compared to those CU and with MCI. Consistent with our previous study,^15^ suspected dementia was also associated with the status of being divorced. The living arrangements observed in this cohort highlight the social importance of multi-generational households in Africa, which provide a supportive environment for older adults. Younger family members often care for the elderly, fostering intergenerational bonds and offering assistance during illness. ^30^ In contrast, individuals with dementia in Western countries often reside in private homes with caregivers, assisted living facilities, nursing homes, or other community-based settings.^31^ In Congo, most individuals with dementia live in multi-generational homes without formal or professional assistance. Those with suspected dementia required more help with complex daily activities compared to the CU and MCI groups.

Cognitively, this cohort was assessed using validated screening and cognitive tests, most of which demonstrated excellent psychometric properties—such as high internal consistency measured by Cronbach’s alpha. However, three tests—the African Visuospatial Memory Test, the African Proverb Test, and the African Emotion Test—showed weaker psychometric performance.

Medically, while SDOH affect adults globally, our cohort faced fewer SDOH-related challenges compared to some Western populations.^32^ Poverty plays a significant role in shaping health outcomes, cognitive aging, and dementia risk in this cohort. Unlike many global cohorts, where few participants have experienced war (e.g., in the U.S., only 6.1% of adults are veterans),^33^ nearly half of our participants (43.5%) reported wartime experience. According to Sixsmith et al.,^34^ such experiences may foster adaptability, independence, and resilience—traits reflected in participants’ resilience scores. However, Burnes et al.^35^ highlighted the detrimental effects of stress and trauma, which can accelerate neurological damage and increase the risk of cognitive decline and dementia.

Regarding neuropsychiatric symptoms (NPS), prior studies have categorized them into three clusters affective symptoms (depression, anxiety, apathy, sleep, and appetite disturbances), agitation symptoms (disinhibition, agitation, irritability, motor disturbances, and euphoria), and psychotic symptoms (hallucinations and delusions).^36^ In our cohort, mood symptoms were the most prevalent, followed by agitation, with few or no psychotic symptoms. As in previous studies, NPS were more frequent among individuals with suspected dementia than in those CU or with MCI.^37^

The prevalence of tobacco smoking in this cohort aligns more closely with rates in USA^38^ and Sub-Saharan Africa (SSA), where adult smoking prevalence averages 12.7%.^39^ A meta-analysis found that smoking rates among adults aged 65 and older in SSA range from 1.8% (Zambia) to 25.8% (Sierra Leone), with men smoking more than women.^40^ Sleep disorders, another dementia risk factor, were also prevalent. While approximately 40% of older adults globally report poor sleep, individuals with AD experience more sleep disturbances due to comorbidities.^41^ In our cohort, sleep disorder prevalence ranged from 27% to 59%, consistent with global estimates of 14% to 59%.^41^ A meta-analysis suggested that sleep disturbances may predict future dementia risk. Insomnia was linked to increased AD incidence, while sleep-disordered breathing was associated with all-cause dementia, AD, and vascular dementia.^42^ Low body weight and BMI in this cohort may reflect poor appetite and confounding factors such as smoking. Weight loss is commonly observed in dementia patients, though the underlying mechanisms remain unclear.^43,44^

Consistent with existing literature, our findings show a high prevalence of vascular conditions, particularly hypertension, which plays a significant role in cognitive decline and dementia.^45^ Additionally, the cohort’s estimated glomerular filtration rate (eGFR) indicated mildly reduced kidney function, which may affect biomarker clearance and plasma concentration levels. Therefore, eGFR should be considered in statistical analyses involving biomarkers.

This study confirms recent research identifying Kinshasa—a megacity with over 17 million residents, projected to reach 35 million by 2040—as the “most polluted major city in the world.”^46^ The city’s air quality exceeds the World Health Organization (WHO) guideline of 5 μg/m³ for PM2.5, contributing to numerous health issues and an estimated 32,000 deaths annually in the Democratic Republic of the Congo.^47^ The primary contributors to poor air quality include widespread biomass burning, elevated emissions from household charcoal stoves, and persistent environmental contaminants such as arsenic, lead, DDT derivatives, and PCBs.^46^ In 2019, McFarlane and colleagues^48^ deployed a five-node PurpleAir monitoring network and reported a calibrated annual average PM2.5 concentration of 43.5 μg/m³ in Kinshasa. Our findings align with earlier studies showing that PM2.5 levels peak during the dry season—June, July, and August—when most of our data were collected. ^47^ Therefore, air pollution should be recognized as a significant and modifiable risk factor for dementia in the Democratic Republic of Congo.

In sum, the 2024 Lancet Commission outlined 12 modifiable risk factors for dementia across the life course, grouped into lifestyle factors (physical inactivity, smoking, ^45,49^excessive alcohol consumption), health conditions (obesity, hypertension, diabetes, depression), environmental factors (air pollution, traumatic brain injury), and cognitive/social factors (low education, social isolation, hearing impairment).^3^ Our cohort reported several of these risk factors, particularly health condition (hypertension and depression), cognitive/social factors (low education and hearing impairment), and environmental (air pollution). Most of these modifiable dementia risk factors are late-life ones which can be prevented.

Our cohort study demonstrates several notable strengths. First, it highlights the feasibility of establishing a research cohort in a low- and middle-income country (LMIC) with limited resources. Despite cultural sensitivities—such as the belief that blood donation threatens life— we successfully collected a wide range of biological samples, including CSF, blood, urine, saliva, and feces. These samples will enable pioneering multi-omics research on SSA populations. Second, the study offers valuable insights into both modifiable and non-modifiable factors associated with cognitive aging and dementia in Africa. These findings have the potential to inform targeted prevention strategies and public health interventions across SSA. Third, the project has fostered interdisciplinary collaboration among laboratory technicians, psychologists, physicians, and other specialists, strengthening local research capacity. It also contributes to public awareness in the DRC, where dementia is often misunderstood as witchcraft, spiritual affliction, or mental illness. Fourth, Kinshasa is emerging as a regional center for dementia research and clinical care, with potential influence extending beyond the DRC to Central African and Francophone countries. The prospective design of the study further enhances its scientific value by allowing for the establishment of temporal relationships between exposures and outcomes, supporting causal inference rather than simple associations. This design improves data accuracy, pre-analytic rigor, and control over confounding variables, thereby increasing internal validity and enabling a deeper understanding of cognitive decline.

Nonetheless, the study has limitations. It is currently restricted to Kinshasa, and future expansion to other provinces is necessary to improve ethnic diversity, enhance external validity, and broaden dementia awareness nationwide. Financial constraints delayed the acquisition of a −80°C freezer and prevented the collection of neuroimaging data. Additionally, a few cognitive tests demonstrated poor to acceptable internal consistency, which may affect measurement reliability. As with most longitudinal studies, participant attrition due to dropout and mortality is expected, potentially reducing sample size and statistical power. Maintaining participant engagement over time has also posed challenges.

Future plans of our cohort study include the possibility to conduct a clinical trial based on the POINTER or FINGER models, incorporating both therapeutic and lifestyle interventions to reduce and prevent dementia in SSA. We also intend to expand the cohort of EVCD-RDC beyond Kinshasa to examine incidence, mortality rates, and survival outcomes of dementia in the DRC. Improving the psychometric properties of the three less reliable cognitive tests is a priority. Given the high prevalence of wartime trauma, infectious diseases, vascular conditions, sleep disorders, and air pollution in our sample, we aim to investigate their contributions to dementia risk. Finally, we plan to develop cognitive and plasma biomarker profiles across the spectrum of cognitive aging, dementia stages, and subtypes in the DRC.

## ACKNOWLEDGEMENTS

There are no acknowledgements.

## AUTHOR CONTRIBUTIONS

JI: Conceptualization; Methodology; Software; Validation; Formal analysis; Investigation; Resources; Data Curation; Writing – original draft; Writing – review & editing; Visualization; Supervision; Project administration; Funding acquisition. CO: Writing – review & editing, Writing – original draft, preparation of figures. SSP: Writing – review & editing, Writing – original draft. MS: Writing – review & editing, Writing – original draft. GG: Writing – review & editing. EE: Writing – review & editing. JP: Writing – review & editing. TT: Writing – review & editing. JK: Writing – review & editing. IK: Writing – review & editing. FS: Writing – review & editing. BM: Writing – review & editing. JT: Writing – review & editing. FB: Writing – review & editing. SM: Writing – review & editing. LM: Writing – review & editing. JB: Writing – review & editing. CT: Writing – review & editing. KT: Writing – review & editing. AG: Writing – review & editing. AA: Writing; Data analysis, Investigation, preparation of figures – review & editing.

## DATA AVAILABILITY

The raw data supporting the conclusions of this article will be made available by the authors, without undue reservation.

## ETHICS STATEMENT

The studies involving humans were approved by University of Kinshasa and Emory University. The studies were conducted in accordance with the local legislation and institutional requirements. The participants provided their written informed consent to participate in this study

## REFERENCES

1. Nichols E, Steinmetz JD, Vollset SE, et al. Estimation of the global prevalence of dementia in 2019 and forecasted prevalence in 2050: an analysis for the Global Burden of Disease Study 2019. thelancet.comE Nichols, JD Steinmetz, SE Vollset, K Fukutaki, J Chalek, F Abd-Allah, A AbdoliThe Lancet Public Health, 2022•thelancet.com 2022; 7: e105–e125.

2. Wimo A, Guerchet M, Ali GC, et al. The worldwide costs of dementia 2015 and comparisons with 2010. Wiley Online LibraryA Wimo, M Guerchet, GC Ali, YT Wu, AM Prina, B Winblad, L Jönsson, Z Liu, M PrinceAlzheimer’s & Dementia, 2017•Wiley Online Library 2017; 13: 1–7.

3. Livingston G, Huntley J, Sommerlad A, et al. Dementia prevention, intervention, and care: 2020 report of the Lancet Commission. thelancet.comG Livingston, J Huntley, A Sommerlad, D Ames, C Ballard, S Banerjee, C Brayne, A BurnsThe lancet, 2020•thelancet.com, https://www.thelancet.com/article/S0140-6736(20)30367-6/fulltext?utm_source=google&utm_medium=sem&utm_campaign=W360_PMax_HearingTest&utm_plasource=banner/&utm_source=google&utm_medium=sem&utm_name=dec_23_traffic&utm_campaign=W360_PMax_HearingTest&utm_plasource=banner&utm_id=HP%20Social%20Media%20Organic%20Post (accessed 1 September 2025).

4. Kingston A, Jagger C. Review of methodologies of cohort studies of older people. academic.oup.comA Kingston, C JaggerAge and Ageing, 2018•academic.oup.com 2018; 47: 215–219.

5. Seshadri S, Wolf PA, Beiser A, et al. Lifetime risk of dementia and Alzheimer’s disease: the impact of mortality on risk estimates in the Framingham Study. neurology.orgS Seshadri, PA Wolf, A Beiser, R Au, K McNulty, R White, RB D’agostinoNeurology, 1997•neurology.org 1997; 49: 1498–1504.

6. Singh-Manoux A, Dugravot A, Fournier A, et al. Trajectories of depressive symptoms before diagnosis of dementia: a 28-year follow-up study. jamanetwork.com 2017; 74: 712–718.

7. Prince M, Acosta D, Ferri CP, et al. Dementia incidence and mortality in middle-income countries, and associations with indicators of cognitive reserve: a 10/66 Dementia Research Group population. thelancet.comM Prince, D Acosta, CP Ferri, M Guerra, Y Huang, JJL Rodriguez, A Salas, AL SosaThe Lancet, 2012•thelancet.com 2012; 380: 50–58.

8. Lima-Costa MF, De Andrade FB, Souza PRB De, et al. The Brazilian longitudinal study of aging (ELSI-Brazil): objectives and design. academic.oup.comMF Lima-Costa, FB de Andrade, PRB Souza, AL Neri, YAO Duarte, E Castro-CostaAmerican journal of epidemiology, 2018•academic.oup.com 2018; 187: 1345–1353.

9. Ogunniyi A, Baiyewu O, Gureje O, et al. Epidemiology of dementia in Nigeria: results from the Indianapolis–Ibadan study. Wiley Online LibraryA Ogunniyi, O Baiyewu, O Gureje, KS Hall, F Unverzagt, SH Siu, S Gao, M FarlowEuropean Journal of Neurology, 2000•Wiley Online Library 2000; 7: 485–490.

10. De Jager CA, Msemburi W, Pepper K, et al. Dementia prevalence in a rural region of South Africa: a cross-sectional community study. journals.sagepub.comCA de Jager, W Msemburi, K Pepper, MI CombrinckJournal of Alzheimer’s Disease, 2017•journals.sagepub.com 2017; 60: 1087–1096.

11. Guerchet M, Mayston R, Lloyd-Sherlock P, et al. Dementia in sub-Saharan Africa: Challenges and opportunities., https://unilim.hal.science/hal-03495416/document (2017, accessed 1 September 2025).

12. Samba H, Guerchet M, Ndamba-Bandzouzi B, et al. Dementia-associated mortality and its predictors among older adults in sub-Saharan Africa: results from a 2-year follow-up in Congo (the EPIDEMCA-FU study). academic.oup.comH Samba, M Guerchet, B Ndamba-Bandzouzi, P Mbelesso, P Lacroix, JF DartiguesAge and ageing, 2016•academic.oup.com 2016; 45: 680–687.

13. Mbelesso P, Tabo A, Guerchet M, et al. Epidemiology of dementia in elderly people in the third district of Bangui (Central African Republic). Bulletin de la société de 2012; 105: 388–395.

14. Guerchet M, Houinato D, Paraiso MN, et al. Cognitive impairment and dementia in elderly people living in rural Benin, west Africa. karger.comM Guerchet, D Houinato, MN Paraiso, N Von Ahsen, P Nubukpo, M Otto, JP ClémentDementia and geriatric cognitive disorders, 2009•karger.com 2009; 27: 34–41.

15. Ikanga J, Patel SS, Roberts BR, et al. Association of plasma biomarkers with cognitive function in persons with dementia and cognitively healthy in the Democratic Republic of Congo. Alzheimer’s and Dementia: Diagnosis, Assessment and Disease Monitoring; 15. Epub ahead of print 1 October 2023. DOI: 10.1002/DAD2.12496.

16. Nkouonlack CD, Njamnshi WY, Angwafor SA, et al. Dementia and cognitive impairment in French-speaking Sub-Saharan Africa: a comprehensive review on moving out of the shadows of neglect. Epub ahead of print 4 May 2023. DOI: 10.21203/RS.3.RS-2887319/V1.

17. Sachs BC, Latham LA, Bateman JR, et al. Feasibility of remote administration of the uniform data set-version 3 for assessment of older adults with mild cognitive impairment and Alzheimer’s disease. journals.aai.orgBC Sachs, LA Latham, JR Bateman, MJ Cleveland, MA Espeland, E Fischer, SA GaussoinArchives of Clinical Neuropsychology, 2024•journals.aai.org 2024; 39: 635–643.

18. Porto MF, Benitez-Agudelo JC, Aguirre-Acevedo DC, et al. Diagnostic accuracy of the UDS 3.0 neuropsychological battery in a cohort with Alzheimer’s disease in Colombia. Taylor & FrancisMF Porto, JC Benitez-Agudelo, DC Aguirre-Acevedo, E Barceló-Martinez, RF AllegriApplied Neuropsychology: Adult, 2022•Taylor & Francis 2022; 29: 1543–1551.

19. Manly JJ, Jones RN, Langa KM, et al. Estimating the prevalence of dementia and mild cognitive impairment in the US: the 2016 health and retirement study harmonized cognitive assessment protocol. jamanetwork.com 2022; 79: 1242–1249.

20. Janelidze S, Barthélemy NR, He Y, et al. Mitigating the associations of kidney dysfunction with blood biomarkers of Alzheimer disease by using phosphorylated tau to total tau ratios. jamanetwork.com 2023; 80: 516–522.

21. Bukabau JB, Yayo E, Gnionsahé A, et al. Performance of creatinine-or cystatin C–based equations to estimate glomerular filtration rate in sub-Saharan African populations. ElsevierJB Bukabau, E Yayo, A Gnionsahe, D Monnet, H Pottel, E Cavalier, A Nkodila, JRR MakuloKidney international, 2019•Elsevier 2019; 95: 1181–1189.

22. Bukabau JB, Sumaili EK, Cavalier E, et al. Performance of glomerular filtration rate estimation equations in Congolese healthy adults: the inopportunity of the ethnic correction. journals.plos.orgJB Bukabau, EK Sumaili, E Cavalier, H Pottel, B Kifakiou, A Nkodila, JRR Makulo, VM MokoliPloS one, 2018•journals.plos.org; 13. Epub ahead of print 1 March 2018. DOI: 10.1371/JOURNAL.PONE.0193384.

23. Gavito-Covarrubias D, Ramírez-Díaz I, Guzmán-Linares J, et al. Epigenetic mechanisms of particulate matter exposure: air pollution and hazards on human health. Front Genet; 14. Epub ahead of print 2023. DOI: 10.3389/FGENE.2023.1306600/FULL.

24. Münzel T, Gori T, Al-Kindi S, et al. Effects of gaseous and solid constituents of air pollution on endothelial function. academic.oup.comT Münzel, T Gori, S Al-Kindi, J Deanfield, J Lelieveld, A Daiber, S RajagopalanEuropean heart journal, 2018•academic.oup.com 2018; 39: 3543–3550.

25. Sun Q, Ren X, Sun Z, et al. The critical role of epigenetic mechanism in PM2.5-induced cardiovascular diseases. SpringerQ Sun, X Ren, Z Sun, J DuanGenes and Environment, 2021•Springer; 43. Epub ahead of print 1 December 2021. DOI: 10.1186/S41021-021-00219-W.

26. Harris PA, Taylor R, Thielke R, et al. Research electronic data capture (REDCap)—a metadata-driven methodology and workflow process for providing translational research informatics support. ElsevierPA Harris, R Taylor, R Thielke, J Payne, N Gonzalez, JG CondeJournal of biomedical informatics, 2009•Elsevier 2009; 42: 377–381.

27. Harris PA, Taylor R, Minor BL, et al. The REDCap consortium: building an international community of software platform partners. ElsevierPA Harris, R Taylor, BL Minor, V Elliott, M Fernandez, L O’Neal, L McLeod, G DelacquaJournal of biomedical informatics, 2019•Elsevier; 95. Epub ahead of print 1 July 2019. DOI: 10.1016/J.JBI.2019.103208.

28. Kinshasa, Urban Pulse of the Congo | National Geographic, https://www.nationalgeographic.com/magazine/article/kinshasa-congo (accessed 1 September 2025).

29. Bassil DT, Farrell MT, Wagner RG, et al. Cohort Profile Update: Cognition and dementia in the Health and Aging in Africa Longitudinal Study of an INDEPTH community in South Africa (HAALSI dementia). academic.oup.comDT Bassil, MT Farrell, RG Wagner, AM Brickman, MM Glymour, KM Langa, JJ ManlyInternational journal of epidemiology, 2022•academic.oup.com 2022; 51: E217–E226.

30. Gouttefarde P, Gay E, Guyot J, et al. The shifts in intergenerational relations in Cameroon and their potential impact on the health of older adults. gh.bmj.comP Gouttefarde, E Gay, J Guyot, O Kamdem, A Socpa, G Tchundem, C Dupré, C NkenfouBMJ Global Health, 2024•gh.bmj.com 2024; 9: 14678.

31. Leverton M, Pui Kin Kor P. Supporting people with dementia to live at home. SpringerM Leverton, P Pui Kin KorBMC geriatrics, 2023•Springer; 23. Epub ahead of print 1 December 2023. DOI: 10.1186/S12877-023-04389-W.

32. Carmichael AE, Lennon NH, Qualters JR. Analysis of social determinants of health and individual factors found in health equity frameworks: Applications to injury research. Elsevier 2023; 87: 508–518.

33. Who are America’s veterans? | USAFacts, https://usafacts.org/articles/who-are-the-nations-veterans/ (accessed 1 September 2025).

34. Sixsmith J, Sixsmith A, Callender M, et al. Wartime experiences and their implications for the everyday lives of older people. cambridge.orgJ Sixsmith, A Sixsmith, M Callender, S CorrAgeing & Society, 2014•cambridge.org 2014; 34: 1457–1481.

35. Burnes DPR, Burnette D. Broadening the etiological discourse on Alzheimer’s disease to include trauma and posttraumatic stress disorder as psychosocial risk factors. Elsevier 2013; 27: 218–224.

36. Therapy TL-AR&, 2020 undefined. Neuropsychiatric symptoms in cognitively normal older persons, and the association with Alzheimer’s and non-Alzheimer’s dementia. SpringerTM LiewAlzheimer’s Research & Therapy, 2020•Springer 2020; 12: 35.

37. Eikelboom WS, Van Den Berg E, Singleton EH, et al. Neuropsychiatric and cognitive symptoms across the Alzheimer disease clinical spectrum: cross-sectional and longitudinal associations. neurology.orgWS Eikelboom, E van den Berg, EH Singleton, SJ Baart, M Coesmans, AE LeeuwisNeurology, 2021•neurology.org 2021; 97: E1276–E1287.

38. Cornelius ME, Loretan CG, Jamal A, et al. Tobacco product use among adults–United States, 2021. cdc.govME CorneliusMMWR Morbidity and mortality weekly report, 2023•cdc.gov 2023; 72: 475–483.

39. Belete H, Mekonen T, Connor JP, et al. Tobacco smoking in Sub Saharan Africa: A systematic review and meta analysis. Wiley Online LibraryH Belete, T Mekonen, JP Connor, G Chan, L Hides, J LeungDrug and Alcohol Review, 2025•Wiley Online Library 2025; 44: 1079–1091.

40. Brathwaite R, Addo J, Smeeth L, et al. A systematic review of tobacco smoking prevalence and description of tobacco control strategies in sub-Saharan African countries; 2007 to 2014. journals.plos.orgR Brathwaite, J Addo, L Smeeth, K LockPloS one, 2015•journals.plos.org 2015; 10: 132401.

41. Bombois S, Derambure P, Pasquier F, et al. Sleep disorders in aging and dementia. SpringerS Bombois, P Derambure, F Pasquier, C MonacaThe journal of nutrition, health & aging, 2010•Springer 2010; 14: 212–217.

42. Shi L, Chen SJ, Ma MY, et al. Sleep disturbances increase the risk of dementia: a systematic review and meta-analysis. ElsevierL Shi, SJ Chen, MY Ma, YP Bao, Y Han, YM Wang, J Shi, MV Vitiello, L LuSleep medicine reviews, 2018•Elsevier 2018; 40: 4–16.

43. Luchsinger JA, Patel B, Tang MX, et al. Body mass index, dementia, and mortality in the elderly. Elsevier 2008; 12: 127–131.

44. Li J, Joshi P, Ang TFA, et al. Mid-to late-life body mass index and dementia risk: 38 years of follow-up of the Framingham Study. academic.oup.comJ Li, P Joshi, TFA Ang, C Liu, S Auerbach, S Devine, R AuAmerican Journal of Epidemiology, 2021•academic.oup.com 2021; 190: 2503–2510.

45. Gottesman RF, Egle M, Groechel RC, et al. Blood pressure and the brain: the conundrum of hypertension and dementia. academic.oup.comRF Gottesman, M Egle, RC Groechel, A MughalCardiovascular Research, 2024•academic.oup.com 2024; 120: 2360–2372.

46. Air Quality Spotlight: Kinshasa, DRC most polluted major city, https://www.iqair.com/ca-fr/newsroom/air-quality-spotlight-biomass-burning-and-pollution-in-kinshasa-democratic-republic-of-the-congo (accessed 20 September 2025).

47. New State of Global Air Special Report on air quality and health in Africa | Health Effects Institute, https://www.healtheffects.org/announcements/new-state-global-air-special-report-air-quality-and-health-africa (accessed 1 September 2025).

48. McFarlane C, Isevulambire PK, Lumbuenamo RS, et al. First Measurements of Ambient PM2.5 in Kinshasa, Democratic Republic of Congo and Brazzaville, Republic of Congo Using Field-calibrated Low-cost Sensors. SpringerC McFarlane, PK Isevulambire, RS Lumbuenamo, AME Ndinga, R Dhammapala, X JinAerosol and Air Quality Research, 2021•Springer; 21. Epub ahead of print 2021. DOI: 10.4209/AAQR.200619.

49. Ge Y. Vascular Contributions to Healthy Aging and Dementia. Aging Dis 2024; 15: 1432.

