## Supplementary material for "Cohort profile: A community-based prospective cohort study of Alzheimer’s disease and related dementias in the Democratic Republic of Congo": Table 1

**Table 1. Tests and questionnaires of the cohort study**

| **Tests** | **Instruments** | **Adapted** | **In Initial Assessment** | **In Follow-up Assessment** |
| --- | --- | --- | --- | --- |
| ***UDS-3*** | A1, A2, A3, A4, A5, B1, B2, B3, B4, C1, C2, D1, & D2 | We adapted some words and expressions to fit Congolese context. | Yes | No |
| ***Screening*** | MOCA | We changed the cube to bed. | Yes | Yes |
|  | CDR | No | Yes | Yes |
|  | IDEA | No | Yes | Yes |
| ***Cognitive -African Neuropsychology Battery -Short Version (ANB-SV)*** | Sequential Movements | No | Yes | Yes |
|  | Tandem Movement | No | Yes | Yes |
|  | Triple loops | No | Yes | Yes |
|  | Mancala | No | Yes | Yes |
|  | African Market Test (AMT) | No | Yes | Yes |
|  | Phonemic Fluency Letter M | We changed letter L to M in both French and Lingala | Yes | Yes |
|  | Semantic Fluency -Food naming | No | Yes | Yes |
|  | African Naming Test (ANT) | We selected 12 items (3 from each of the 4 categories) | Yes | Yes |
|  | African List Memory Test (ALMT) | We removed the interference list of words | Yes | Yes |
|  | African Visuospatial Memory Test (AVMT) | We removed the interference designs | Yes | .  Yes |
|  | African Story Memory Test (ASMT) | We reduced the story to 25 important details. | Yes | Yes |
|  | African Faces Perception Test (AFPT) | We selected 10 out of 20 faces. | Yes | Yes |
|  | African Emotion Recognition Test (AERT) | We used an old Congolese adult face who imitated the 6 basic emotions | Yes | Yes |
|  | African Proverb Test (APT) | We selected only 5 proverbs and used only Congolese proverbs familiar to participants. | Yes | Yes |
|  | African Card Game Total | We selected 4 games out of 7 games. | Yes | Yes |
| ***Intelligence*** | Vocabulary | We translated words into French and Lingala. | Yes | Yes |
|  | Matrix Reasoning | No | Yes | Yes |
| ***Questionnaires*** | Social Determinants of Health | No | Yes | No |
|  | Test of Resilience | We adapted some words and expressions to fit Congolese context. | Yes | No |
|  | The Multidimensional Poverty Measure (MPM) | We adapted some words and expressions to fit Congolese context. | Yes | No |
|  | Air Pollution | No | Yes | Yes |
|  | Modifiable Risk Factors for Cardiovascular Health | No | Yes | Yes |
|  | Geriatric Evaluation of Cumulative Disease | No | Yes | Yes |
|  | Functional Literacy | No | Yes | No |
