## Supplementary material for "Cohort profile: A community-based prospective cohort study of Alzheimer’s disease and related dementias in the Democratic Republic of Congo": Table 2

*Table 2. Characteristics of the Cohort Stratified by Neurological Status*

| **Variables,**  Mean (SD) | **Overall**  (n=506) | **CU**  (n=70; 14%) | **MCI**  (n=136; 27%) | **AD**  (n=120; 24%) | **OD**  (n=180; 36%) | **p-value** |
| --- | --- | --- | --- | --- | --- | --- |
| **Demographics and Social Determinants of Health** | | | | | | |
| **Age** | 73.4 (5.9) | 71.6 (5.1) | 72.6 (5.4) | 75.2 (6.7) | 73.7 (5.7) | 0.13 |
| **Female*** | 300 (59%) | 33 (47%) | 80 (59%) | 64 (53%) | 123 (68%) | 0.11 |
| **Linguistic Group*** |  |  |  |  |  | 0.97 |
| Lingala | 82 (16%) | 12 (17%) | 22 (16%) | 17 (14%) | 31 (17%) |  |
| Kikongo | 165 (33%) | 28 (40%) | 41 (30%) | 39 (33%) | 57 (32%) |  |
| Swahili | 30 (5.9%) | 4 (5.7%) | 11 (8.1%) | 7 (5.8%) | 8 (4.4%) |  |
| Tshiluba | 74 (15%) | 11 (16%) | 21 (15%) | 15 (13%) | 27 (15%) |  |
| Other | 155 (31%) | 15 (21%) | 41 (30%) | 42 (35%) | 57 (32%) |  |
| **Ethnic Group*** |  |  |  |  |  | 0.63 |
| Bantu | 497 (98%) | 70 (100%) | 132 (97%) | 118 (98%) | 177 (98%) |  |
| Nilotique | 3 (0.59%) | 0 (0%) | 1 (0.74%) | 1 (0.83%) | 1 (0.56%) |  |
| Soudanique | 4 (0.79%) | 0 (0%) | 2 (1.5%) | 0 (0%) | 2 (1.1%) |  |
| Other | 2 (0.40%) | 0 (0%) | 1 (0.74%) | 1 (0.83%) | 0 (0%) |  |
| **Education, Years** | 9.7 (4.8) | 12.1 (4.1) | 10.7 (4.3) | 8.8 (5.0)^†^ | 8.7 (5.0)^†^ | 0.013 |
| **Marital Status*** |  |  |  |  |  | 0.074 |
| Married | 216 (43%) | 39 (56%) | 70 (52%) | 44 (37%) | 63 (35%) |  |
| Widowed | 247 (49%) | 24 (34%) | 55 (40%) | 61 (51%) | 107 (59%) |  |
| Divorced | 16 (3.2%) | 4 (5.7%) | 4 (2.9%) | 5 (4.2%) | 3 (1.7%) |  |
| Separated | 8 (1.6%) | 0 (0%) | 2 (1.5%) | 3 (2.5%) | 3 (1.7%) |  |
| Single | 14 (2.8%) | 2 (2.9%) | 4 (2.9%) | 4 (3.3%) | 4 (2.2%) |  |
| Living with a Partner | 2 (0.40%) | 0 (0%) | 0 (0%) | 2 (1.7%) | 0 (0%) |  |
| Unknown | 3 (0.59%) | 1 (1.4%) | 1 (0.74%) | 1 (0.83%) | 0 (0%) |  |
| **Living Arrangement*** | | |  |  |  | 0.0006 |
| Lives Alone | 13 (2.6%) | 1 (1.4%) | 4 (2.9%) | 5 (4.2%) | 3 (1.7%) |  |
| Lives with Spouse/Partner | 196 (39%) | 38 (54%) | 60 (44%) | 33 (28%) | 65 (36%) |  |
| Lives with Relative/Friend | 194 (38%) | 24 (34%) | 49 (36%) | 46 (38%) | 75 (42%) |  |
| Lives with Caregiver | 1 (0.20%) | 0 (0%) | 0 (0%) | 1 (0.83%) | 0 (0%) |  |
| Lives with a Group | 98 (19%) | 7 (10%) | 22 (16%) | 32 (27%) | 37 (21%) |  |
| Lives in a Group Home | 2 (0.40%) | 0 (0%) | 0 (0%) | 2 (1.7%) | 0 (0%) |  |
| Unknown | 2 (0.40%) | 0 (0%) | 0 (0%) | 1 (0.83%) | 0 (0%) |  |
| **Level of Independence*** | | |  |  |  | <.0001 |
| Able to Live Independently | 311 (62%) | 59 (84%) | 100 (74%) | 56 (47%) | 96 (53%) |  |
| Needs Help with Complex Activities | 150 (30%) | 10 (14%) | 30 (22%) | 42 (35%)^†^ | 68 (38%)^†^ |  |
| Needs Help with Basic Activities | 34 (6.7%) | 0 (0%) | 3 (2.2%) | 18 (15%) | 13 (7.2%) |  |
| Completely Dependent | 9 (1.8%) | 1 (1.4%) | 2 (1.5%) | 3 (2.5%) | 3 (1.7%) |  |
| Unknown | 2 (0.40%) | 0 (0%) | 1 (0.74%) | 1 (0.83%) | 0 (0%) |  |
| **Type of Residence*** | | |  |  |  | 0.64 |
| Private Residence | 501 (99%) | 70 (100%) | 134 (99%) | 118 (98%) | 179 (99%) |  |
| Assisted Living Facility | 2 (0.40%) | 0 (0%) | 0 (0%) | 1 (0.83%) | 1 (0.56%) |  |
| Unknown | 3 (0.59%) | 0 (0%) | 2 (1.5%) | 1 (0.83%) | 0 (0%) |  |

*Results presented as n(%).

^†^Indicates significant pairwise association between the neurological status category and cognitively normal (the reference group).

Note: Abbreviations: CU=Cognitively Unimpaired, MCI = Mild Cognitive Impairment, AD = Alzheimer’s Disease, OD = Other Dementia.
