## Supplementary material for "Cohort profile: A community-based prospective cohort study of Alzheimer’s disease and related dementias in the Democratic Republic of Congo": Table 3

*Table 3. Medical Profile of Cohort, Stratified by Neurological Status*

| **Variables,**  Mean (SD) | **Overall**  (n=506) | **CU**  (n=70; 14%) | **MCI**  (n=136; 27%) | **AD**  (n=120; 24%) | **OD**  (n=180; 36%) | **p-value** |
| --- | --- | --- | --- | --- | --- | --- |
| **Physical Evaluation** | | | | | | |
| Height, cm | 162.6 (8.6) | 165.4 (7.4) | 165.0 (8.5) | 161.6 (9.3) | 160.4 (8.0) | 0.11 |
| Weight, kg | 69.6 (18.5) | 78.3 (25.4) | 73.2 (19.5) | 68.6 (17.9) | 64.0 (12.8)^†^ | 0.025 |
| BMI, kg/m^2^ | 25.9 (5.1) | 26.3 (4.5) | 27.2 (5.2) | 24.9 (4.6) | 26.2 (6.9) | 0.10 |
| Systolic BP, mmHg | 141.6 (21.9) | 147.9 (21.7) | 137.0 (21.1)^†^ | 138.4 (22.8)^†^ | 144.3 (21.2) | 0.009 |
| Diastolic BP, mmHg | 85.5 (12.9) | 86.3 (11.6) | 84.8 (13.0) | 84.4 (13.0) | 86.2 (13.5) | 0.69 |
| Stroke History* | 11 (2.2%) | 0 (0%) | 2 (1.5%) | 3 (2.5%) | 6 (3.3%) | 0.41 |
| Heart Rate, bpm | 72.4 (10.7) | 72.1 (9.0) | 72.2 (10.9) | 71.3 (10.7) | 73.4 (11.3) | 0.79 |
| Wears Glasses* | 188 (37%) | 36 (51%) | 55 (40%) | 35 (29%) | 62 (34%) | 0.16 |
| Normal Hearing* | 472 (93%) | 65 (93%) | 130 (96%) | 106 (88%) | 171 (95%) | 0.23 |
| Hypertension |  |  |  |  |  | 0.69 |
| Good | 43 (8.5%) | 7 (26%) | 15 (11%) | 13 (11%) | 8 (4.4%) |  |
| Intermediate | 64 (13%) | 8 (11%) | 16 (12%) | 23 (19%) | 17 (9.4%) |  |
| Poor | 106 (21%) | 17 (24%) | 25 (18%) | 36 (30%) | 28 (16%) |  |
| Unknown | 293 (58%) | 38 (54%) | 80 (59%) | 48 (40%) | 127 (71%) |  |
| Aware of Hypertension | 134 (27%) | 20 (29%) | 34 (25%) | 42 (35%) | 38 (21%) | 0.080 |
| Years with Hypertension* | 9.6 (7.0) | 7.0 (8.5) | 8.0 (4.2) | 9.5 (6.9) | 11.5 (7.8) | 0.82 |
| In Hypertension Treatment | 82 (16%) | 15 (21%) | 20 (15%) | 21 (18%) | 26 (14%) | 0.045 |
| Hypertension Controlled | 81 (16%) | 15 (21%) | 22 (16%) | 27 (23%) | 17 (9.4%) | 0.93 |
| **Abnormal Neurological Exam Results*** | 107 (21%) | 15 (21%) | 13 (9.6%) | 24 (20%) | 55 (31%) | 0.0004 |
| **Functional Evaluation Total** | 42.7 (10.8) | 42.3 (11.6) | 45.0 (9.0) | 42.8 (12.0) | 40.8 (10.7) | 0.15 |
| **Cumulative Evaluation of Disease – Mild to Severe Impairment*** | | | | | | |
| Heart | 21 (4.2%) | 3 (4.3%) | 9 (6.6%) | 2 (1.7%) | 7 (3.9%) | 0.11 |
| Vascular | 86 (17%) | 15 (21%) | 18 (13%) | 25 (21%) | 28 (16%) | 0.46 |
| Hematopoietic | 4 (0.79%) | 0 (0%) | 0 (0%) | 2 (1.7%) | 2 (1.1%) | 0.54 |
| Respiratory | 15 (3.0%) | 0 (0%) | 2 (1.5%) | 5 (4.2%) | 8 (4.4%) | 0.089 |
| Eyes, Ears, Nose, Throat | 192 (38%) | 26 (37%) | 45 (33%) | 50 (42%) | 71 (39%) | 0.84 |
| Upper Gastrointestinal | 61 (12%) | 9 (13%) | 14 (10%) | 18 (15%) | 20 (11%) | 0.34 |
| Lower Gastrointestinal | 29 (5.7%) | 4 (5.7%) | 2 (1.5%) | 11 (9.2%) | 12 (6.7%) | 0.046 |
| Renal | 3 (0.59%) | 0 (0%) | 0 (0%) | 2 (1.7%) | 1 (0.56%) | 0.42 |
| Liver | 3 (0.59%) | 1 (1.4%) | 0 (0%) | 1 (0.83%) | 1 (0.56%) | 0.42 |
| Urogenital | 54 (11%) | 11 (16%) | 9 (6.6%) | 18 (15%) | 16 (8.9%) | 0.11 |
| Musculoskeletal | 108 (21%) | 18 (26%) | 21 (15%) | 37 (31%) | 32 (18%) | 0.022 |
| Endocrine | 21 (4.2%) | 3 (4.3%) | 4 (2.9%) | 5 (4.2%) | 9 (5.0%) | 0.42 |
| Neurological | 30 (5.9%) | 4 (5.7%) | 2 (1.5%) | 10 (8.3%) | 14 (7.8%) | 0.030 |
| Psychiatric | 3 (0.59%) | 1 (1.4%) | 0 (0%) | 1 (0.83%) | 1 (0.56%) | 0.45 |
| **Creatinine, mg/dL** |  |  |  |  |  |  |
| Mean (SD) | 0.90 (0.30) | 0.90 (0.30) | 0.90 (0.30) | 0.90 (0.40) | 0.90 (0.30) | 0.75 |
| Median (IQR) | 0.85 (0.30) | 0.85 (0.35) | 0.84 (0.34) | 0.87 (0.28) | 0.84 (0.28) | 0.62 |
| **eGFR Labs** |  |  |  |  |  |  |
| MDRD | 71.9 (16.2) | 74.4 (16.7) | 72.0 (15.3) | 69.7 (16.7) | 72.3 (16.2) | 0.30 |

*Results presented as n(%).

^†^Indicates significant pairwise association between the neurological status category and cognitively normal (the reference group).
