## Supplementary material for "Cohort profile: A community-based prospective cohort study of Alzheimer’s disease and related dementias in the Democratic Republic of Congo": Table 4

*Table 4. Neuropsychiatric Profiles of the Cohort Stratified by Neurological Status*

| **Variables,**  Mean (SD) | **Overall**  (n=506) | **CU**  (n=70; 14%) | **MCI**  (n=136; 27%) | **AD**  (n=120; 24%) | **OD**  (n=180; 36%) | **p-value** |
| --- | --- | --- | --- | --- | --- | --- |
| **Experience of War*** | 220 (44%) | 26 (37%) | 67 (49%) | 55 (46%) | 72 (40%) | 0.19 |
| **Traumatic Event in Last 10 Years*** | 300 (59%) | 37 (53%) | 79 (58%) | 70 (58%) | 114 (63%) | 0.54 |
| **Smoker*** | 60 (12%) | 8 (11%) | 18 (13%) | 15 (13%) | 19 (11%) | 0.80 |
| **# of Years Smoking** | 1.4 (6.2) | 2.0 (6.7) | 2.0 (7.0) | 1.3 (6.8) | 0.8 (4.8) | 0.13 |
| **Frequency of Alcohol Intake*** | | |  |  |  | 0.14 |
| Never | 4 (0.79%) | 0 (0%) | 1 (0.74%) | 0 (0%) | 3 (1.7%) |  |
| Once a month | 105 (21%) | 18 (26%) | 31 (23%) | 24 (20%) | 32 (18%) |  |
| 2-4 times a month | 31 (6.1%) | 5 (7.1%) | 16 (12%) | 3 (2.5%) | 7 (3.9%) |  |
| 2-3 times per week | 22 (4.4%) | 4 (5.7%) | 11 (8.1%) | 4 (3.3%) | 3 (1.7%) |  |
| 4+ times a week | 15 (3.0%) | 4 (5.7%) | 4 (2.9%) | 3 (2.5%) | 4 (2.2%) |  |
| Unknown | 329 (65%) | 39 (56%) | 73 (54%) | 86 (72%) | 131 (73%) |  |
| **Sleep Disorder*** | 155 (31%) | 24 (34%) | 36 (27%) | 40 (33%) | 55 (31%) | 0.47 |
| **SDOH Total** | 6.2 (2.7) | 7.5 (2.3) | 6.4 (2.7) | 5.9 (2.8) | 5.7 (2.8)^†^ | <.0001 |
| **Resiliency Total** | 19.7 (3.5) | 21.3 (2.4) | 21.0 (2.6) | 18.0 (4.0)^†^ | 19.2 (3.6) | <.0001 |
| **Dimension of Poverty Total** | 6.1 (2.7) | 4.7 (2.7) | 5.8 (2.6) | 6.8 (2.4)^†^ | 6.5 (2.8)^†^ | <.0001 |

*Results presented as n(%)

^†^Indicates significant pairwise association between the neurological status category and cognitively normal (the reference group).
