## Supplementary material for "Cohort profile: A community-based prospective cohort study of Alzheimer’s disease and related dementias in the Democratic Republic of Congo": Table 5

*Table 5. Internal Consistency of Screening and Cognitive Tests*

| **Test** | **# of Items** | **Total** | **Cronbach’s Alpha** | **Corrected**  **Item-Total Correlations^a^** | **Mean Inter-Item Correlations** |
| --- | --- | --- | --- | --- | --- |
| MoCA | 28 | 30 | 0.85 | (0.24, 0.62) | 0.18 |
| CDR | 6 | 6 | 0.90 | (0.624, 0.83) | 0.62 |
| IDEA | 11 | 11 | 0.91 | (0.57, 0.74) | 0.51 |
| African Market Test | 19 | 20 | 0.91 | (0.20, 0.72) | 0.34 |
| ALMT^b^ | 12 | 12 | 0.82 | (0.61, 0.75) | 0.62 |
| AVMT^c^ | 4 | 20 | 0.78 | (0.50, 0.69) | 0.51 |
| ASMT^b^ | 25 | 25 | 0.91 | (.76, 0.88) | 0.78 |
| Emotion Recognition | 6 | 6 | 0.44 | (0.06, 0.38) | 0.12 |
| Proverb Test | 5 | 10 | 0.74 | (0.37, 0.61) | 0.36 |
| African Card Game | 25 | 25 | - | - | - |

^a^ Range of corrected item-total correlations is reported.

^b^ Internal consistency, Corrected item- total correlations, and mean inter-item correlations are reported for the three initial encoding items.

^c^ Internal consistency, Corrected item- total correlations, and mean inter-item correlations are reported for the four figure copy items.

Note: Abbreviations: ALMT=African List Memory Test (ALMT), AVMT=African Visuospatial Memory Test, ASMT=African Story Memory Test.
