## Supplementary material for "Cohort profile: A community-based prospective cohort study of Alzheimer’s disease and related dementias in the Democratic Republic of Congo": Table 6

*Table 6. Cohort Cognitive Assessments Stratified by Neurological Status*

| **Variables,**  Mean (SD) | **Overall**  (n=506) | **CU**  (n=70; 14%) | **MCI**  (n=136; 27%) | **AD**  (n=120; 24%) | **OD**  (n=180; 36%) | **p-value** |
| --- | --- | --- | --- | --- | --- | --- |
| MCA | 19.4 (5.1) | 24.2 (3.6) | 22.0 (2.9) | 15.8 (5.3)^†^ | 18.2 (4.2)^†^ | <.0001 |
| CDR | 3.3 (3.5) | 1.3 (1.5) | 2.0 (1.9) | 5.5 (4.5)^†^ | 3.5 (3.4)^†^ | <.0001 |
| **Psychomotricity** |  |  |  |  |  |  |
| Sequential Movements-Dominant Hand | 7.8 (3.2) | 8.7 (2.7) | 8.7 (2.3) | 6.8 (3.7) | 7.5 (3.4) | <.0001 |
| Sequential Movements-Non-Dominant Hand | 7.9 (3.5) | 8.2 (3.3) | 9.1 (2.5) | 6.8 (4.0) | 7.6 (3.6) | <.0001 |
| Tandem Movement | 11.0 (2.8) | 11.9 (0.6) | 11.6 (2.1) | 9.8 (4.1)^†^ | 11.0 (2.8) | <.0001 |
| **Attention** |  |  |  |  |  |  |
| Mancala-Total Points | 4.1 (2.0) | 5.6 (1.4) | 4.8 (1.6) | 2.9 (2.0)^†^ | 3.7 (1.9)^†^ | <.0001 |
| Mancala-Total Correct | 27.5 (13.8) | 37.7 (10.1) | 34.2 (11.1) | 18.9 (13.1)^†^ | 24.2 (12.5)^†^ | <.0001 |
| Mental Reversal-Total Points | 4.2 (2.2) | 5.9 (1.9) | 5.1 (1.8) | 2.7 (2.2)^†^ | 3.9 (1.9)^†^ | <.0001 |
| Mental Reversal-Total Correct | 26.0 (15.3) | 39.1 (12.5) | 32.5 (12.8)^†^ | 15.0 (13.3)^†^ | 23.4 (13.2)^†^ | <.0001 |
| **Working Memory** |  |  |  |  |  |  |
| African Market Test | 14.3 (10.6) | 17.9 (20.2) | 14.3 (5.3) | 12.4 (5.8) | 12.9 (6.3) | 0.024 |
| **Ideational Praxis** | 4.4 (0.9) | 4.7 (0.5) | 4.5 (0.7) | 4.1 (1.1)^†^ | 4.3 (0.9)^†^ | <.0001 |
| **Fluency** |  |  |  |  |  |  |
| Phonemic | 5.5 (3.9) | 8.2 (4.1) | 6.0 (3.8)^†^ | 4.0 (3.5)^†^ | 5.0 (3.6)^†^ | <.0001 |
| Semantic | 10.9 (4.4) | 14.0 (3.7) | 11.7 (4.1)^†^ | 8.9 (4.0)^†^ | 10.3 (4.4)^†^ | <.0001 |
| **Learning and Memory** | | |  |  |  |  |
| ALMT Trial 1 | 4.8 (1.7) | 5.9 (1.5) | 5.2 (1.4)^†^ | 3.8 (1.6)^†^ | 4.8 (1.7)^†^ | <.0001 |
| ALMT Trial 2 | 6.4 (1.8) | 7.6 (1.7) | 6.9 (1.6)^†^ | 5.4 (1.8)^†^ | 6.2 (1.7)^†^ | <.0001 |
| ALMT Trial 3 | 7.2 (2.1) | 9.0 (1.4) | 7.8 (1.5)^†^ | 5.9 (2.1)^†^ | 6.9 (2.0)^†^ | <.0001 |
| ALMT Delayed Recall | 4.9 (2.7) | 7.7 (1.7) | 5.9 (2.0)^†^ | 2.3 (2.1)^†^ | 4.9 (2.2)^†^ | <.0001 |
| ALMT Force Choice | 10.8 (2.1) | 11.7 (0.8) | 11.4 (1.3) | 9.3 (3.1)^†^ | 10.9 (1.7) | <.0001 |
| AVMT Trial 1 | 3.6 (3.2) | 6.3 (3.7) | 4.9 (3.3) | 2.0 (2.2)^†^ | 2.6 (2.2)^†^ | <.0001 |
| AVMT Trial 2 | 4.9 (3.9) | 8.4 (3.8) | 6.8 (3.8) | 2.8 (2.7)^†^ | 3.6 (3.0)^†^ | <.0001 |
| AVMT Trial 3 | 6.0 (4.6) | 10.9 (3.9) | 8.2 (4.3)^†^ | 3.4 (3.1)^†^ | 4.2 (3.5)^†^ | <.0001 |
| AVMT Delayed Recall | 5.7 (4.5) | 11.1 (3.9) | 7.7 (4.2)^†^ | 3.0 (3.2)^†^ | 4.0 (3.2)^†^ | <.0001 |
| AVMT Forced Choice | 3.9 (0.5) | 4.0 (0.0) | 3.9 (0.3) | 3.7 (0.8)^†^ | 3.8 (0.6) | <.0001 |
| ASMT Trial 1 | 6.7 (4.2) | 10.1 (3.9) | 8.3 (3.8) | 3.9 (3.6)^†^ | 6.0 (3.5)^†^ | <.0001 |
| ASMT Trial 2 | 9.4 (5.1) | 14.3 (3.5) | 11.6 (4.2)^†^ | 5.9 (4.3)^†^ | 8.1 (4.4)^†^ | <.0001 |
| ASMT Trial 3 | 10.6 (5.5) | 16. 0 (3.6) | 13.2 (4.5)^†^ | 6.9 (4.7)^†^ | 9.1 (4.8)^†^ | <.0001 |
| ASMT Delayed Recall | 8.8 (5.4) | 14.4 (4.0) | 11.1 (4.3)^†^ | 4.9 (4.3)^†^ | 7.6 (4.6)^†^ | <.0001 |
| ASMT Forced Choice | 4.0 (1.3) | 4.8 (0.4) | 4.6 (0.9) | 3.3 (1.5)^†^ | 3.8 (1.2)^†^ | <.0001 |
| **Executive Functions** | | |  |  |  |  |
| Proverb Test | 2.6 (2.5) | 5.3 (2.2) | 3.0 (2.3)^†^ | 1.8 (2.4)^†^ | 1.7 (2.0)^†^ | <.0001 |
| African Card Game | 12.5 (5.3) | 16.4 (4.5) | 13.7 (4.5)^†^ | 10.1 (4.7)^†^ | 11.7 (5.4)^†^ | <.0001 |
| **Visuospatial Perception** | 5.7 (2.5) | 7.2 (1.7) | 6.3 (1.9) | 4.8 (3.3)^†^ | 5.3 (2.1)^†^ | <.0001 |
| **Emotion Recognition** | 3.5 (1.2) | 3.9 (1.1) | 3.9 (1.0) | 2.9 (1.4)^†^ | 3.6 (1.2) | <.0001 |
| **Intelligence** |  |  |  |  |  |  |
| Verbal Intelligence | 21.8 (12.6) | 29.8 (12.7) | 23.9 (11.6) | 18.4 (12.7)^†^ | 19.4 (11.6)^†^ | <.0001 |
| Non-Verbal Matrix Test | 5.6 (3.9) | 9.3 (4.6) | 6.4 (3.5)^†^ | 3.9 (3.5)^†^ | 4.7 (3.2)^†^ | <.0001 |
| Full Scall IQ-2 | 71.3 (15.5) | 83.5 (17.6) | 73.6 (14.1)^†^ | 66.8 (14.3)^†^ | 68.1 (13.8)^†^ | <.0001 |

^†^Indicates significant pairwise association between the neurological status category and cognitively normal (the reference group).
