## Supplementary material for "Cohort profile: A community-based prospective cohort study of Alzheimer’s disease and related dementias in the Democratic Republic of Congo": Table 7

*Table 7. Environmental Characteristics and Air Pollution Measures Across Diagnostic Groups*

| **Variables**  Mean (SD) | **Overall**  **(N=156)** | **CU**  **(N=22)** | **MCI**  **(N=42)** | **AD**  **(N=48)** | **OD**  **(N=44)** |
| --- | --- | --- | --- | --- | --- |
| PM 1 | 90.73 (73.17) | 81.29 (53.63) | 101.97 (111.91) | 79.98 (43.20) | 98.14 (64.16) |
| PM 2.5 | 102.32 (104.21) | 82.38 (59.12) | 109.04 (116.47) | 85.93 (45.97) | 125.53 (147.24) |
| PM 10 | 98.41 (77.36) | 90.36 (59.36) | 109.62 (116.25) | 87.11 (46.70) | 105.99 (69.29) |
| Barometric Pressure | 980.30 (11.58) | 977.46 (17.05) | 982.01 (2.48) | 978.13 (16.87) | 982.51 (2.07) |
| COV | 0.13 (0.06) | 0.11 (0.06) | 0.13 (0.06) | 0.13 (0.06) | 0.14 (0.06) |
| Temperature | 25.59 (2.56) | 25.60 (2.65) | 25.61 (2.31) | 25.42 (2.87) | 25.77 (2.47) |
| Humidity | 58.10 (6.67) | 58.87 (6.10) | 57.86 (6.70) | 58.45 (7.45) | 57.54 (6.17) |
| Altitude | 278.51 (101.70) | 303.37 (148.90) | 263.44 (21.19) | 297.63 (148.68) | 259.15 (17.66) |
